# Three-Dimensional Echocardiographic Evaluation of Longitudinal and Non-Longitudinal Components of Right Ventricular Contraction Results from the World Alliance of Societies of Echocardiography Study

**DOI:** 10.1101/2023.04.20.23288902

**Authors:** Juan I. Cotella, Attila Kovacs, Karima Addetia, Alexandra Fabian, Federico M. Asch, Roberto M. Lang, the WASE Investigators

**Author notes:** **Correspondence to:** Roberto M Lang, MD, The University of Chicago Medicine, 5758 S. Maryland Avenue, MC 9067, DCAM 5509, Chicago, IL 60637.

## Abstract

**Background:** Right ventricular (RV) functional assessment is mainly limited to its longitudinal component. However, due to the complex orientation of the myofibers, the RV contraction involves coordinated motion along multiple planes. Recently developed 3-dimensional echocardiography (3DE) software has enabled the separate assessment of the non-longitudinal components of RV systolic function and their relative contribution to RV performance. The aims of this study were 1) to establish normal values for 3D-derived longitudinal, radial, and anteroposterior RV ejection fraction (LEF, REF, AEF respectively) and their relative contributions to global RVEF, 2) to calculate 3D RV strain normal values and, 3) to determine sex, age and race related differences in these parameters in a large group of normal subjects (WASE study)

**Methods:** 1043 healthy adult subjects prospectively enrolled at 17 centers in 15 countries were used in this study. 3DE RV wide-angle datasets were analyzed to generate a 3D mesh model of the RV cavity (TomTec). Then, dedicated software (ReVISION) was used to analyze RV motion along the three main anatomical planes and the ejection fraction (EF) values corresponding to each plane were identified as LEF, REF, and AEF. Relative contributions were determined by dividing each EF component by the global RVEF. RV strain analysis included longitudinal, circumferential, and global area strains (GLS, GCS and GAS, respectively). Results were categorized by sex, age (18-40, 41-65 and >65 years), and race.

**Results:** Absolute REF, AEF, LEF and global RVEF were higher in women than in men (p < .001). With aging, both sexes exhibited a decline in all the determinants of longitudinal shortening (p <.001). In elderly women, the lower global RVEF was partially compensated by an increase in radial shortening. Both Black men and women showed lower RVEF, and GAS values compared to White and Asian subjects of the same sex (p < .001). Black men showed significantly higher REF/RVEF and lower LEF/RVEF compared to Asian and White men. These differences in RV contraction patterns across races were not present in women.

**Conclusion:** 3DE evaluation of the non-longitudinal components of RV contraction provides additional information regarding RV physiology, including sex, age and race - related differences in RV contraction patterns that may prove useful in disease states involving the RV.

## INTRODUCTION

Right ventricular (RV) function has a prognostic role in several cardiovascular diseases ^1–4^. Since these conditions are frequently associated with RV remodeling and dysfunction, a comprehensive assessment of this chamber has become pivotal in cardiovascular evaluation. In daily clinical practice, assessment of RV morphology and systolic function is predominantly performed using transthoracic echocardiography (TTE) and is based on the interpretation of images obtained from multiple transducer positions with two-dimensional (2D) measurements, such as: basal and mid-ventricular diameters, fractional area change (FAC), tricuspid annular plane systolic excursion (TAPSE), tissue Doppler tricuspid annular velocity, and RV free-wall strain (RV FWS).

However, the right ventricle has a complex, crescent-shaped anatomy, combined with an intricate contraction pattern. Due to the different orientation of its subepicardial (circumferential) and subendocardial (longitudinal) myocardial fibers ^5^, RV contraction occurs along the longitudinal, radial and anteroposterior directions. As a result, the use of simple 2D echocardiographic (2DE) techniques might be inadequate to accurately evaluate RV systolic performance, as they provide information limited to the in-plane components of RV contraction.

Three-dimensional echocardiography (3DE) allows the evaluation of the entire right ventricle from a single acquisition. Through the generation, manipulation and decomposition of 3DE dynamic surface rendering models of the right ventricle, software has been recently developed to enable the separate assessment of the longitudinal and non-longitudinal components of RV systolic function and quantify their relative contributions to global RV performance ^6^. However, little is known about what these components look like in the normal population, and therefore there is no reference basis to detect abnormalities.

Accordingly, the aims of this study were 1) to establish normal values for longitudinal, radial, and anteroposterior motion components of RV deformation and determine their relative contributions to global RV performance in a large group of normal subjects, and 2) to examine sex, age, and race related differences in these normal values. To achieve this goal, we used the population from the World Alliance Societies of Echocardiography (WASE) Study, which represents the largest collection of normal 2DE and 3DE TTE images.

## METHODS

### Study Design and Population

The WASE Study rationale was described in detail elsewhere ^7, 8^. A comprehensive 2D and 3D TTE examination was performed in all study subjects using commercially available ultrasound imaging systems (GE, Philips, and Siemens). Image acquisition followed a standard protocol established by ASE/EACVI guidelines and data analyzed by the two WASE core laboratories (University of Chicago and MedStar Health Research Institute) ^9^.

### Image Acquisition and Analysis

Wide-angle 3DE full-volume RV datasets were acquired with the patient in the left decubitus position over 4-6 cardiac cycles during suspended respiration from the RV-focused view, while carefully avoiding stitching and drop-out artifacts. Data was digitally stored and analyzed offline at the 3DE Core Lab (University of Chicago) using dedicated vendor-independent software (Image Arena; “4D RV-Function”, TOMTEC, Unterschleissheim, Germany), previously validated against cardiac magnetic resonance (CMR) reference ^9, 10^. 3DE datasets were deemed adequate for analysis if all walls were visible throughout the cardiac cycle. A minimum frame rate of 15 volumes per second was required for the software to perform the analysis.

Auto-segmentation technology with the help of specific manually identified landmarks allowed the 3D RV dataset to be displayed in both the long and short-axis cut-planes with an initial endocardial border contour suggestion. End-diastole was automatically identified as the time point in which the RV cavity was the largest, while end-systole was identified as the time-point when the cavity was the smallest. Afterwards, the user could manually adjust the proposed end-diastolic and end-systolic endocardial border contours to optimize dynamic tracking throughout the cardiac cycle. Once this step was completed, the program generated and rendered a 3D RV surface model.

### Decomposition of the RV motion

Figure 1. illustrates the workflow for the decomposition of RV systolic function into its different contractile components.

**Fig. 1.**
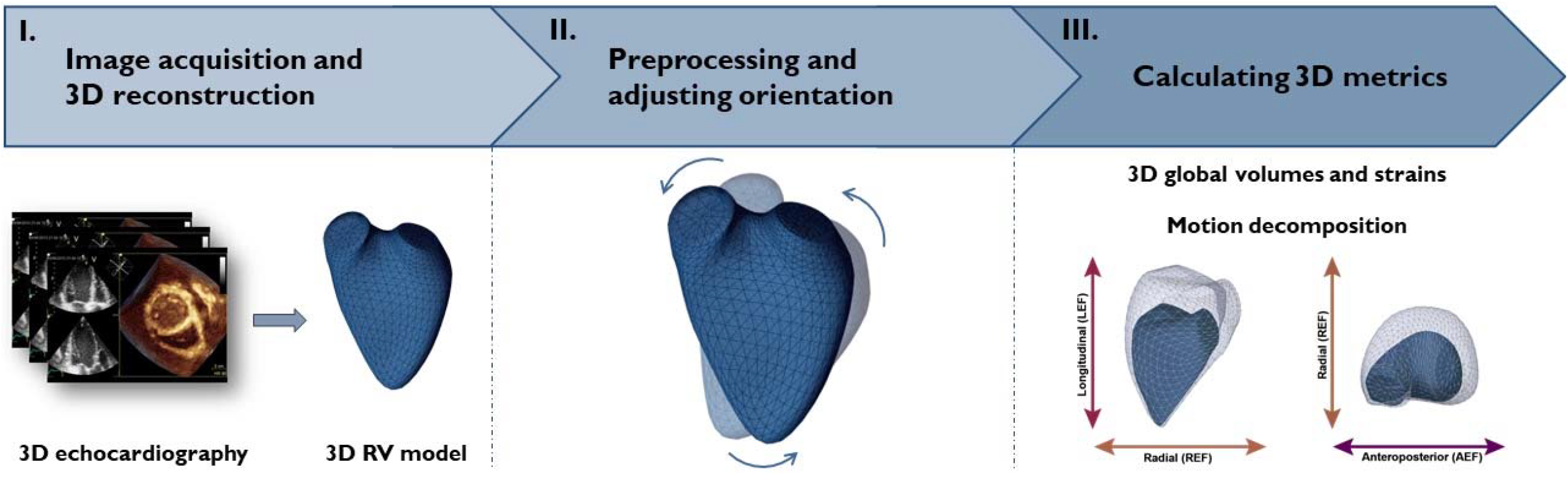
Right Ventricular Contraction Decomposition Workflow

#### 1. 3D reconstruction and mesh model import

First, the 3D RV models (formulated as a series of polygon meshes) were exported volume-by-volume throughout the cardiac cycle from the TOMTEC software. Then, these meshes were imported to ReVISION software (Right VentrIcular Separate wall motIon quantificatiON; Argus Cognitive, Lebanon, New Hampshire) ^6^ used in the next steps.

#### 2. Preprocessing and standardization of the 3D mesh orientation

To standardize the orientation and further decomposition of the RV contraction, the 3D RV models were aligned to a reference mesh by methodology described elsewhere ^11^ aiming to define three anatomically relevant, orthogonal axes of the RV: longitudinal, radial and anteroposterior.

#### 3. Movement decomposition and calculation of 3D RV EF

Initially, 3D RV end-diastolic volume indexed (EDVi) to body surface area (BSA) and global 3D RVEF were calculated. Once the relevant axes were defined, the wall motion of the 3D RV model was decomposed in a vertex-based manner. This step transformed the original 3D polygon mesh model into several series of meshes, each corresponding to a decomposition type, allowing the isolation and independent assessment of the magnitude of longitudinal, radial or anteroposterior motions.

By decomposing the model’s motion along the predefined orthogonal axes, the software was capable to separately quantify the RV volume changes along each direction. The volumes of the models related to only one motion direction were calculated at each time frame using the signed tetrahedron method ^6^. The corresponding EF value for each axis was defined as longitudinal, radial and anteroposterior EF (LEF, REF and AEF, respectively). The relative and individual contribution of LEF, REF and AEF to the global RV performance, were calculated as ratios, dividing each axis EF by the global RVEF (LEF/RVEF, REF/RVEF and AEF/RVEF respectively). Importantly, the absolute volume change generated by the software corresponds to the aggregated contribution of the three motion components. This composition is not additive, and consequently, the sum of the decomposed volume changes is not equal to the global RV volume change ^12^.

#### 4. Calculation of 3D RV Strain

Finally, 3D RV global longitudinal strain (GLS) and global circumferential strain (GCS), were calculated by the changes in predefined longitudinal and circumferential contour lengths referenced to the corresponding end-diastolic contour length.

Briefly, to obtain 3D RV GLS, 45 longitudinally oriented contours were generated by connecting the apex and predefined vertices of the base through specific equidistant vertices at the middle section by fitting geodesic lines (the shortest path between two points on a curved surface). This method ensures that the longitudes are evenly distributed on the surface of the mesh.

Ultimately, the quantification of the change in the length of each RV longitudinal contour throughout the entire cardiac cycle allowed the calculation of the 3D RV GLS.

For the calculation of 3D RV GCS, the pulmonary and tricuspid annular planes were excluded. Then, 15 circumferential contours were created by slicing horizontally the 3D RV mesh model at equal distances along the longitudinal axis. After generating a set of circumferential contours, we were able to quantify the change in their positions at later time instants and to provide 3D RV GCS values.

Additionally, 3D RV global area strain (GAS) was defined as the percentage change in the endocardial area of the 3D models. Further details about the analytical method of ReVISION software have been documented elsewhere ^11^.

### Statistical Analysis

All data are presented as the mean ± standard deviation (SD). Group differences were evaluated using ANOVA and unpaired two-tailed student’s t-tests. Statistical significance was defined as p<0.05. Normal ranges for each parameter were defined as upper and lower limits using 2.5th and 97.5th percentile values from the relevant group. This is in accordance with the definition of “normal” as falling within 95% of the normal population, with the remaining 5% being distributed equally between the two tails of the distribution, irrespective of whether it is Gaussian. Pearson or Spearman correlation tests were applied, as appropriate. In addition, multivariable linear regression analyses were performed to identify independent associations between the decomposed RV metrics with demographic, anthropometric, basic hemodynamic, and LV functional parameters. In order to add race, we dichotomized our cohort accordingly, analyzing White (yes/no), Black (yes/no) and Asian (yes/no) races as separate variables in each model to avoid collinearity.

## RESULTS

Out of the initial cohort of 2262 subjects from the WASE Study, 2007 had 3D RV datasets available in the format suitable for measurement. Of these, 1043 (52%) had adequate image quality. Exclusion criteria included: 1) low frame rates (<15 volumes per second), 2) presence of stitch artifact, 3) excessive drop-out of the anterior RV free wall, and 4) incomplete data capture with the lateral free-wall or the RV apex being cut-off from the pyramidal volume, suggesting that data was collected from the 4-chamber instead of the RV-focused view. **Table 1** shows the basic anthropometric and demographic data of the study population. Men and women were equally distributed. Most of the population was white (47%) followed by Asians (36%). Subjects were distributed in six sub-groups according to age and sex: 18-40 years (234 men, 199 women), 41-65 years (194 men, 197 women) and >65 years (105 men and 114 women). Upper and lower limits of normal (ULN and LLN, respectively) for global and direction-specific RV functional parameters are provided in **Table 2**.

**Table 1.** Basic anthropometric and demographic data in the total study population and separately in men and women.

|  | All subjects<br>n=1043 | Men<br>n=533 | Women<br>n=510 | P-value |
| --- | --- | --- | --- | --- |
| **Age (years)** | 46.8±16.6 | 46.2±16.4 | 47.4±16.8 | 0.245 |
| **Heart rate (bpm)** | 64.6±10.5 | 63.5±10.3 | 65.7±10.5 | **0.001** |
| **BSA, m<sup>2</sup>** | 1.78±0.21 | 1.89±0.19 | 1.66±0.17 | **<0.001** |
| **Race** |  |  |  |  |
| • **Asian** | 375 (36%) | 206 (39%) | 169 (33%) | 0.064 |
| • **Black** | 142 (14%) | 69 (13%) | 73 (14%) | 0.519 |
| • **White** | 488 (47%) | 235 (44%) | 253 (50%) | 0.074 |
| • **Other (Mixed and other)** | 38 (4%) | 23 (4%) | 15 (3%) | 0.236 |

ml: milliliters, bpm: beats per minute

**Table 2.** Upper and lower limits of normality for the novel RV parameters in men and women

|  | All subjects<br>n=1043 | Men<br>n=533 | Women<br>n=510 |
| --- | --- | --- | --- |
|  | **LLN to ULN** | **LLN to ULN** | **LLN to ULN** |
| **RV EDVi (ml)** | 43.7 to 123.1 | 47.9 to 132.5 | 41.7 to 110.6 |
| **RV EF (%)** | 44.7 to 67.1 | 44.1 to 65.7 | 45.8 to 68.6 |
| **RV GCS** | -13.4 to -31.3 | -13.9 to -30.9 | -12.8 to -31.6 |
| **RV GLS** | -13.2 to -27.4 | -13.4 to -26.9 | -12.5 to -27.9 |
| **RV GAS** | -28.4 to -48.9 | -28.5 to -47.6 | -28.4 to -49.6 |
| **REF (%)** | 15.3 to 42.7 | 15.0 to 41.8 | 15.7 to 43.1 |
| **REF/RVEF** | 0.30 to 0.73 | 0.29 to 0.73 | 0.30 to 0.73 |
| **AEF (%)** | 14.1 to 37.2 | 13.7 to 35.3 | 14.2 to 37.8 |
| **AEF/RVEF** | 0.28 to 0.61 | 0.28 to 0.60 | 0.28 to 0.61 |
| **LEF (%)** | 11.1 to 33.4 | 11.7 to 31.7 | 11.0 to 35.0 |
| **LEF/RVEF** | 0.22 to 0.56 | 0.23 to 0.55 | 0.22 to 0.57 |

ml: milliliters, LLN: lower limit normal (2.5th percentile), ULN: upper limit normal (97.5th percentile), RV EDVi: right ventricular end-diastolic volume index, RV EF: right ventricular ejection fraction, RV GCS: right ventricular global circumferential strain, RV GLS: right ventricular global longitudinal strain, RV GAS: right ventricular global area strain, REF: radial ejection fraction, AEF: anteroposterior ejection fraction, LEF: longitudinal ejection fraction

### 1. Sex-related differences

**Table 3** shows the 3D RV morphological and functional parameters of the total study population and separated by sex. RVEDVi was significantly higher in men than in women (p <0.001). Global RVEF and absolute REF, AEF, LEF values were higher in women than in men (p <0.001). However, the relative contribution of each of the individual components to the global RVEF was similar in both sexes. While GLS and GAS were significantly higher in women than in men, GCS showed no differences between sexes.

**Table 3.** Sex-related differences in 3D RV morphological and functional parameters

|  | All subjects<br>n=1043 | Men<br>n=533 | Women<br>n=510 | P- value |
| --- | --- | --- | --- | --- |
| **RV EDVi (ml)** | 76.4 ± 20.2 | 82.2 ± 21.4 | 70.5 ± 16.9 | **<0.001** |
| **RV EF (%)** | 55.5 ± 5.8 | 54.5 ± 5.4 | 56.5 ± 6.0 | **<0.001** |
| **RV GCS** | -22.1 ± 4.5 | -21.9 ± 4.3 | -22.3 ± 4.8 | 0.175 |
| **RV GLS** | -19.7 ± 3.7 | -19.4 ± 3.5 | -20.1 ± 3.9 | **0.008** |
| **RV GAS** | -38.0 ± 5.2 | -37.3 ± 4.8 | -38.6 ± 5.6 | **<0.001** |
| **REF (%)** | 29.0 ± 7.0 | 28.3 ± 6.7 | 29.7 ± 7.1 | **0.001** |
| **REF/RVEF** | 0.52 ± 0.11 | 0.52 ± 0.11 | 0.53 ± 0.11 | 0.284 |
| **AEF (%)** | 25.4 ± 5.9 | 24.7 ± 5.7 | 26.1 ± 6.1 | **<0.001** |
| **AEF/RVEF** | 0.45 ± 0.08 | 0.45 ± 0.08 | 0.46 ± 0.08 | 0.188 |
| **LEF (%)** | 21.6 ± 5.7 | 21.0 ± 5.2 | 22.3 ± 6.0 | **<0.001** |
| **LEF/RVEF** | 0.39 ± 0.09 | 0.38 ± 0.08 | 0.39 ± 0.09 | 0.144 |

### 2. Age-related differences

**Table 4** shows the age-related differences in the components of 3D RV systolic function in both men and women. There were no significant differences in RVEDVi between age groups. Both sexes exhibited a decline in longitudinal shortening (i.e., LEF, LEF/RVEF, and GLS, p < .001) with increasing age. In women, this reduction in LEF/RVEF led to a lower global RVEF (p < 0.001), which was incompletely compensated by an increase in REF/RVEF.

**Table 4.** Age-related differences in 3D RV morphological and functional parameters ^a^P < 0.05 vs. 18–40-year age group ^b^P < 0.05 vs. 41–65-year age group ^c^P < 0.05 vs. > 65 years age group y: years, ml: milliliters, ANOVA: analysis of variance, RV EDVi: right ventricular end-diastolic volume index, RV EF: right ventricular ejection fraction, RV GCS: right ventricular global circumferential strain, RV GLS: right ventricular global longitudinal strain, RV GAS: right ventricular global area strain, REF: radial ejection fraction, AEF: anteroposterior ejection fraction, LEF: longitudinal ejection fraction

| Men (n=533) |  |  |  |  | Women (n=510) |  |  |  |
| --- | --- | --- | --- | --- | --- | --- | --- | --- |
|  | 18-40 y<br>n=234 | 41-65 y<br>n=194 | >65 y<br>n=105 | p-ANOVA | 18-40 y<br>n=199 | 41-65 y<br>n=197 | >65 y<br>n=114 | p-ANOVA |
| RV EDVi (ml) | 83.4±19.4 | 80.6±21.9 | 82.3±24.3 | 0.364 | 71.1±17.0 | 70.0±16.5 | 70.0±17.3 | 0.783 |
| RV EF (%) | 54.7±5.5 | 54.7±5.4 | 53.5±5.1 | 0.094 | 56.9±5.9 <sup>c</sup> | 57.1±5.9 <sup>c</sup> | 55.0±6.1 <sup>a,b</sup> | **0.005** |
| RV GCS | -22.0±4.4 | -22.0±4.6 | -21.8±3.5 | 0.914 | -22.4±5.0 | -22.5±4.7 | -21.8±4.6 | 0.281 |
| RV GLS | -20.0±3.4 <sup>c</sup> | -19.5±3.6 <sup>c</sup> | -18.2±3.4 <sup>a,b</sup> | **<0.001** | -20.9± 3.9 <sup>b,c</sup> | -20.1±3.8 <sup>a,c</sup> | -18.6±3.7 <sup>a,b</sup> | **<0.001** |
| RV GAS | -37.7±4.8 <sup>c</sup> | -37.6±4.9 <sup>c</sup> | -36.0±4.2 <sup>a,b</sup> | **0.008** | -39.4±5.6 <sup>c</sup> | -38.9±5.4 <sup>c</sup> | -36.8±5.5 <sup>a,b</sup> | **<0.001** |
| REF (%) | 28.1±6.8 | 28.5±7.0 | 28.6±6.3 | 0.724 | 28.7±7.2 <sup>b,c</sup> | 30.5±7.2 <sup>a</sup> | 30.4±6.6 <sup>a</sup> | **0.021** |
| REF/RVEF | 0.51±0.11 | 0.52±0.11 | 0.53±0.10 | 0.233 | 0.50±0.11 <sup>b,c</sup> | 0.53±0.11 <sup>a</sup> | 0.55±0.1 <sup>a</sup> | **<0.001** |
| AEF (%) | 24.6±5.6 | 25.0±5.8 | 24.4±5.6 | 0.690 | 26.3±6.2 | 26.5±5.9 | 25.0±6.2 | 0.098 |
| AEF/RVEF | 0.45±0.1 | 0.45±0.09 | 0.45±0.1 | 0.699 | 0.46±0.09 | 0.46±0.1 | 0.45±0.1 | 0.668 |
| LEF (%) | 21.8±5.1 <sup>c</sup> | 21.2±5.3 <sup>c</sup> | 18.9±4.9 <sup>a,b</sup> | **<0.001** | 23.5±6.1 <sup>c</sup> | 22.5±5.5 <sup>c</sup> | 19.8±6.1 <sup>a,b</sup> | **<0.001** |
| LEF/RVEF | 0.40±0.1 <sup>c</sup> | 0.39±0.1 <sup>c</sup> | 0.35±0.1 <sup>a,b</sup> | **<0.001** | 0.41±0.09 <sup>b,c</sup> | 0.39±0.1 <sup>a,c</sup> | 0.36±0.1 <sup>a,b</sup> | **<0.001** |

### 3. Race-related differences

BSA and RVEDVi were significantly smaller in Asian compared to black and white subjects (**Table 5**). Both black men and women showed lower RVEF, and GAS values compared to white and Asian subjects of the same sex (p <0.001). Black men showed a distinctive RV mechanical pattern, consistent of significantly higher REF/RVEF and lower LEF/RVEF values, when compared to Asian and white men. White men showed higher LEF/RVEF than both black and Asian men (p <0.001). Of note, these differences in RV contraction patterns across races were not significant in women.

**Table 5.** Race-related differences in 3D RV morphological and functional parameters ^a^P < 0.05 vs. 18–40-year age group ^b^P < 0.05 vs. 41–65-year age group ^c^P < 0.05 vs. > 65 years age group ml: milliliters, ANOVA: analysis of variance, RV EDVi: right ventricular end-diastolic volume index, RV EF: right ventricular ejection fraction, RV GCS: right ventricular global circumferential strain, RV GLS: right ventricular global longitudinal strain, RV GAS: right ventricular global area strain, REF: radial ejection fraction, AEF: anteroposterior ejection fraction, LEF: longitudinal ejection fraction

| Men (n=510) |  |  |  |  | Women (n=495) |  |  |  |
| --- | --- | --- | --- | --- | --- | --- | --- | --- |
|  | White<br>n=235 | Black<br>n=69 | Asian<br>n=206 | p-ANOVA | White<br>n=253 | Black<br>n=73 | Asian<br>n=169 | p-ANOVA |
| RV EDVi (ml) | 85.7±21.6<sup>b,c</sup> | 95.5±23.7<sup>a,c</sup> | 74.4±17<sup>a,b</sup> | **<0.001** | 70.9±16.7<sup>b,c</sup> | 81.2±16.8<sup>a,c</sup> | 65.7±16<sup>a,b</sup> | **<0.001** |
| RV EF (%) | 55.4±5.9<sup>b,c</sup> | 52.4±4.5<sup>a,c</sup> | 54.2±5.0<sup>a,b</sup> | **<0.001** | 57.2±6.1<sup>b</sup> | 54.9±5.1<sup>a</sup> | 56.2±6.1 | **0.010** |
| RV GCS | -22.4±4.4<sup>c</sup> | -21.4±4.1 | -21.5±4.2<sup>a</sup> | **0.034** | -22.5±4.7 | -22.1±4.6 | -22.1±5.1 | 0.627 |
| RV GLS | -20.2±3.5<sup>b,c</sup> | -18.2±3.3<sup>a</sup> | -19.1±3.5<sup>a</sup> | **<0.001** | -20.1±4.0 | -19.2±3.8 | -20.3±3.9 | 0.119 |
| RV GAS | -38.3±5.0<sup>b,c</sup> | -35.2±4.2<sup>a,c</sup> | -37.0±4.5<sup>a,b</sup> | **<0.001** | -38.7±5.6<sup>b</sup> | -37.1±5.0<sup>a,c</sup> | -39.0±5.8<sup>b</sup> | **0.036** |
| REF (%) | 28.1±7.1 | 29.1±5.7 | 28.3±6.7 | 0.598 | 30.3±7.5 | 29.3±6.4 | 29.3±7.0 | 0.261 |
| REF/RVEF | 0.50±0.11<sup>b</sup> | 0.55±0.09<sup>a,c</sup> | 0.52±0.11<sup>b</sup> | **0.004** | 0.53±0.11 | 0.53±0.10 | 0.52±0.11 | 0.656 |
| AEF (%) | 25.5±5.8<sup>b</sup> | 22.5±4.9<sup>a,c</sup> | 24.6±5.7<sup>b</sup> | **0.001** | 26.3±6.1 | 24.9±4.8 | 26.2±6.7 | 0.221 |
| AEF/RVEF | 0.46±0.08<sup>b</sup> | 0.43±0.08<sup>a</sup> | 0.45±0.9 | **0.046** | 0.46±0.08 | 0.45±0.07 | 0.46±0.09 | 0.706 |
| LEF (%) | 22.3±5.2<sup>b,c</sup> | 18.0±4.8<sup>a,c</sup> | 20.6±4.9<sup>a,b</sup> | **<0.001** | 22.8±6.1<sup>b</sup> | 20.3±5.7<sup>a,c</sup> | 22.3±6.1<sup>b</sup> | **0.011** |
| LEF/RVEF | 0.40±0.08<sup>b,c</sup> | 0.34±0.08<sup>a,c</sup> | 0.38±0.1<sup>a,b</sup> | **<0.001** | 0.40±0.09 | 0.37±0.09 | 0.39±0.09 | 0.087 |

### 4. Association with 3D LV functional parameters and independent predictors of the contractile patterns

Correlations between LEF, REF, AEF and basic demographic, anthropometric, hemodynamic and LV functional parameters are summarized in **Supplementary Table 1**. Notably, three-dimensional echo-derived LV EF correlated weakly with 3D RV EF (r=0.161, p<0.001), LEF (r=0.112, p=0.001), and with AEF (r=0.126, p<0.001). 3D LV GLS also correlated weakly with 3D RV EF (r= -0.158, p<0.001), 3D RV GLS (r=0.166, p<0.001) and LEF (r= -0.238, p<0.001).

Results of multivariable linear regression analyses are shown in **Supplementary Tables 2,3 and 4**. Beyond age, sex, and LVEF, white and black races were independent predictors of LEF, whereas sex, LVEF, and white and black races were independent predictors of AEF.

## DISCUSSION

The main findings of this study are: 1) Smaller RVEDVi and higher RV EF values, with significantly higher GLS and GAS contributions are seen in women compared to men; 2) There is a significant decrease in global RV EF in older subjects of both sexes, mainly driven by a reduction in the longitudinal component of RV contraction; 3) Only in women, this reduction of RVEF with increasing age is partially compensated by an increase in REF; 4) The effects of race on RV contraction patterns are particularly significant in men; 5) While black subjects have significantly higher radial and lower longitudinal contributions to global RVEF, white men have higher values of longitudinal and anteroposterior components.

### Rationale for the evaluation of non-longitudinal components of RV contraction

The RV myo-architecture and contraction are extremely complex. While the septum is characterized by oblique longitudinal and spiral fibers, the RV free wall contains predominantly transversal myofibers ^13^. Due to this unique spatial anatomical disposition of myofibers, RV contraction is the result of the combination of three distinctive contractile patterns i.e., shortening along the longitudinal axis, radial contraction of the free wall (“bellows effect”), and anteroposterior shortening as a result of left ventricular (LV) contraction.

However, all 2DE techniques used for the evaluation of RV contraction (i.e. TAPSE and S wave tissue Doppler velocity) provide information on regional longitudinal, rather than global RV function. Although 2DE RV FWS has the advantage of being a more sensitive indicator of subclinical impairment of RV contraction than conventional 2DE measurements ^2, 14, 15^, it has been reported that in situations where RV function is not predominantly affected by reduced longitudinal contraction or where global RV function remains preserved despite decreased longitudinal motion, the sole use of 2DE RV FWS might be insufficient to characterize RV systolic function ^16, 17^.

Whenever examiners are familiar with 3DE imaging and image quality is acceptable, it should be used the evaluation of RV function. This is supported by previous reports showing that 3DE assessment of RV volumes, EF and GLS are feasible and reproducible ^18^, and provides additional prognostic information in patients with diverse cardiac conditions ^19, 20^. Moreover, constant advances and development of matrix-array transducers with higher spatial and temporal resolutions has made possible the development of dedicated software for 3DE deconstruction of RV systolic function in all its motion components. The software used in our study has previously shown excellent intra- and inter-observer reproducibility as well as a robust correlation with currently available echocardiographic software and a modest correlation with CMR ^11, 21^. These findings and the existing evidence about the clinical and prognostic value of the measurements performed in our study, support the feasibility and potential role of this approach in enhancing our understanding of the mechanisms of RV adaptation or maladaptation to pathologic conditions.

### Evidence about the clinical utility of the different components of RV systolic function

The prognostic value of the parameters included in our study has been described by Kitano et al. who showed that 3D RV GCS, GLS and GAS were significantly associated with cardiac death, ventricular tachyarrhythmia, or heart failure hospitalization at 20 months of follow-up in a cohort of patient with diverse cardiac diseases ^21^. Moreover, these findings persisted after adjusting for factors like age, renal function, LVEF and average mitral E/e’. Kaplan-Meier analysis showed that median values of 3D RV GCS, GLS and GAS stratified these patients by survival rates ^21^. Tolvaj et al have described the association between reduced RV GCS and significantly increased risk of mortality, even in scenarios where LV GLS is preserved ^22^.

Interestingly, 3D RV GAS was shown to be a better prognostic marker than 3DE RV GLS or GCS, most likely due to the fact that the information provided by GAS corresponds to an multiplanar evaluation of RV contraction and is not limited to either the longitudinal or circumferential directions as are GLS and GCS ^21^.

It is key to understand that the RV adaptive mechanisms vary according to the underlying pathophysiological process. These different mechanical responses can be identified only through the separate assessment of the multiple components of RV contraction. This approach has allowed to describe distinctive RV mechanical adaptations in different populations ^21, 23–25^, including heart transplant recipients ^26^, congenital cardiomyopathies ^27^, and elite athletes ^28^.

In a large cohort of patients with mildly and moderately reduced LVEF, Surkova et al. demonstrated that despite initially reduced LEF and AEF, REF increased, leading to maintain the global RV performance. Once the contribution of REF decreased, the global RV EF became severely reduced ^29^. Notably, in patients with preserved RV EF (>45%), the AEF component has shown to be a significant and independent predictor of outcome after a median of 6.7 years follow-up ^29^. These findings raise the question about the role of the evaluation of RV contraction beyond the longitudinal shortening in patients with LV dysfunction, considering that identifying the decrease in other motion components could be useful as early markers or RV dysfunction, follow-up parameters or indicators for treatment escalation. Accordingly, in a meta-analysis of more than 1,900 patients, Sayour et al. have elegantly demonstrated that 3D RVEF has a stronger association with adverse cardiopulmonary outcomes than conventional RV functional indices, namely TAPSE, FAC and RV FWS ^30^.

Severe mitral regurgitation (MR) represents one of the most frequent indications for valve surgery, and its progression leads to adaptive (and maladaptive) changes in both left and right ventricles. Due to its broadly recognized prognostic implications, timely diagnosis and appropriate risk stratification represent a major clinical need ^31^. Accordingly, Tokodi et al. reported that in patients with severe primary MR and RV EF > 45%, there was a reduction in REF and a predominant contribution of LEF to the global RV contraction. However, in the immediate post-operative period, this contractile RV pattern inverted, and REF prevailed over LEF. Interestingly, after 6 months, the contribution of both components had equalized ^24^.

These reports reinforce the notion that the impact of different hemodynamic conditions on RV systolic function might be overlooked, if the evaluation of RV contraction is only performed using conventional parameters of longitudinal deformation. The question whether these changes in RV contraction patterns might be useful as surrogates of early RV dysfunction and represent additional criteria for medical/surgical intervention, will require further prospective research in patients with specific pathology.

### Contributions of the current study

Although Lakatos et al. have previously described the contribution of non-longitudinal components of RV contraction in 300 healthy volunteers from a single European and Asian institutions ^12^, our study broadens this analysis to the larger, multicentric and multiethnic WASE cohort, yielding a more statistically sound assessment of the influence of sex, age, and race on these parameters. The LLN and ULN values for 3D RVEDVi and RV EF provided by our study are similar to those reported by the two largest studies of 3DE normative values of this chamber, including Maffessanti et al. ^32^ and Addetia et al. ^33^.

However, current clinical practice lacks RV functional parameters that represent the non-longitudinal shortening of this chamber. The results of our study expands the evidence provided by previous reports, showing that REF and AEF have similar contributions to that of LEF to global RV contraction, and that these components should not be neglected when evaluating the RV function ^12^. Importantly, we were able to show that the RV contraction pattern is independently associated with age, sex, and race highlighting that the established changes across the different subgroups are not just due to anthropometric (BSA) or basic hemodynamic (heart rate, blood pressure) differences. The magnitude of longitudinal and anteroposterior shortenings are coupled with LV function; however, radial shortening is not. This observation is in line with the findings of Surkova et al. who showed that these mechanical directions are deteriorating in parallel with LV systolic dysfunction ^29^.

Our findings agree with those reported in the 2D RV systolic function analysis on the WASE population, which have shown that RV dimensions were larger in men and 2D RV functional parameters were larger in women ^34^. Age-related changes in these 2D parameters were not uniform ^34^ and previous studies using 3DE assessment of RV systolic function have also noted that age is weakly correlated with RVEF ^32^. Notably, we were able to identify an age-related reduction in the longitudinal components of RV contraction in both sexes and a characteristic increase in radial RV contraction in women with age. These observations might be related to an age-related increase in pulmonary pressures and consequent changes in RV myofiber architecture. A preserved RV EF does not preclude the presence of changes in the RV contraction patterns, and the role of REF as an early marker of elevated pulmonary pressures has been proposed ^16^. Muraru et al. have identified LV GLS and PASP as independent predictors of RV longitudinal performance, suggesting that the assessment of the RV adaptation to increased afterload should not be limited to the RV free wall ^35^. Accordingly, our findings support the concept that the evaluation of RV systolic function in individuals > 65 years should not be restricted to its longitudinal motion.

The use of 3DE allowed to show the presence of higher longitudinal and anteroposterior contraction as well as lower radial contributions to global RVEF in Asian population ^12^. Although noticeable differences in 3D RV volumes and EF across ethnic groups have been recently identified ^33^, to the best of our knowledge, this is the first study describing distinctive RV contractile patterns in black and white men, which had been previously overlooked when only using 2DE parameters ^34^. Our findings contribute to expand the understanding of race-related differences in echocardiographic measurements.

## LIMITATIONS

The feasibility of performing a 3D RV analysis in the WASE cohort was slightly above 50% ^33^. It must be acknowledged that 3DE acquisitions of the RV are technically demanding, requiring a high level of sonographer expertise. Although, patients with poor acoustic windows, arrhythmias, large body habitus, obstructive pulmonary disease, or severe chamber dilation are usually excluded from research protocols, these are frequently encountered in daily clinical practice. Low accuracy of automated measurements in right ventricles of different geometry and image quality was also described and should be considered ^36, 37^. Furthermore, in our study there was a relative under-representation of black subjects and individuals from the older age group.

## CONCLUSION

Mechanical adaptations of the right ventricle are complex and are often not restricted to its longitudinal components. The clinical and prognostic utility of the different components of 3DE RV function has been previously reported. Our results reassure this concept through the demonstration of the physiological equivalence of longitudinal and non-longitudinal components of RV contraction and the presence sex, age, and race - related differences in RV contraction patterns. These findings might be useful in considering both physiological and pathological states involving the right ventricle.

## Data Availability

N/A

## Abbreviations

(ASE): American Society of Echocardiography
(BSA): Body surface area
(CMR): Cardiovascular magnetic resonance
(EACVI): European Association of Cardiovascular Imaging
(EDVi): End-diastolic volume index
Ejection Fraction
(FAC): Fractional area change
(AEF): Anteroposterior Ejection Fraction
(EF): Ejection fraction
(GAS): Global area strain
(GCS): Global circumferential strain
(GLS): Global longitudinal strain
(LEF): Longitudinal Ejection Fraction
(MR): Mitral regurgitation
(REF): Radial Ejection Fraction
(RV): Right ventricular
(TAPSE): Tricuspid annular peak systolic excursion
(TTE): Transthoracic Echocardiography
(2DE): Two-dimensional Echocardiography
(3DE): Three-dimensional Echocardiography
(WASE): World Alliance Societies of Echocardiography

**<u>* WASE Investigators</u>**

- **Argentina**: *Aldo D. Prado*, Centro Privado de Cardiologia, Tucumán, Argentina; *Eduardo Filipini*, Universidad Nacional de la Plata, Buenos Aires, Argentina; *Ricardo E. Ronderos*, Instituto Cardiovascular de Buenos Aires, Buenos Aires, Argentina
- **Australia**: *Agatha Kwon and Samantha Hoschke-Edwards*, Heart Care Partners, Queensland, Australia; *Gregory M. Scalia*, Genesis Care, Brisbane, Australia
- **Brazil**: *Tania Regina Afonso and Ana Clara Tude Rodridugues*, Albert Einstein Hospital, Sao Paulo, Brazil
- **Canada**: *Babitha Thampinathan, Maala Sooriyakanthan and Wendy Tsang*, Toronto General Hospital, University of Toronto, Canada
- **China**: *Mei Zhang, Yingbin Wang and Yu Zhang*, Qilu Hospital of Shandong University, Jinan, China; *Tiangang Zhu and Zhilong Wang*, Peking University People’s Hospital, Beijing, China; *Lixue Yin and Shuang Li*, Sichuan Provincial People’s Hospital, Sichuan, China
- **India:** *R. Alagesan*, Madras Medical College, Chennai, India; *S. Balasubramanian*, Madurai Medical College, Madurai, India; *R*.*V*.*A. Ananth and Vivekanandan Amuthan*,Jeyalakshmi Heart Center, Madurai, India; *Manish Bansal*, Medanta Heart Institute, Medanta, Haryana, India; *Ravi R. Kasliwal*, Medanta Heart Institute, Gurgaon, Haryana, India
- **Iran:** *Azin Alizadehasl*, Rajaie Cardiovascular Medical Center, IUMS, Tehran, Iran; *Anita Sadeghpour*, Rajaie Cardiovascular Medical and Research Center, Tehran, Islamic Republic of Iran
- **Italy**: *Luigi Badano and Denisa Muraru*, University of Milano-Bicocca, and Istituto Auxologico Italiano, IRCCS, Milan, Italy; *Eduardo Bossone, Davide Di Vece, Rodolfo Citro and Michele Bellino*, University of Salerno, Salerno, Italy
- **Japan**: *Tomoko Nakao, Takayuki Kawata, Megumi Hirokawa, Naoko Sawada and Masao Daimon*, The University of Tokyo, Tokyo, Japan; *Yousuke Nabeshima and Masaki Takeuchi*, University of Occupational and Environmental Health, Kitakyushu, Japan
- **Republic of Korea**: *Hye Rim Yun, Seung Woo Park and Ji-won Hwang*, Samsung Medical Center, Seoul, Republic of Korea
- **Mexico**: *Pedro Gutierrez Fajardo*, Hospitales Mac Bernardette, Guadalajara, Mexico
- **Nigeria**: *Kofo O. Ogunyankin*, First Cardiology Consultants Hospital, Lagos, Nigeria
- **Phillipines**: *Edwin S. Tucay*, Philippine Heart Center, Quezon City, Philippines
- **United Kingdom**: *Mark J. Monaghan*, King’s College Hospital, London, United Kingdom
- **United States**: *James N. Kirkpatrick*, University of Washington, Seattle, WA; *Tatsuya Miyoshi*, MedStar Health Research Institute, Washington, DC

